# Menopausal hormone therapy at age 45 to 60 years old, future dementia or cognitive decline: Systematic review and meta-analysis

**DOI:** 10.64898/2026.04.20.26351058

**Authors:** Sum Yuet Rainbow Law, Naaheed Mukadam, Nelsan Pourhadi, Amirah Chaudry, Anastasia Shiakalli, Umraj Rai, Gill Livingston, Professor of Psychiatry of Older People

## Abstract

**Objective:** To examine whether menopausal women who initiate systemic menopausal hormone therapy (MHT) around menopause (45-60 years old) have a different risk of developing dementia than those not taking MHT.

**Design:** Systematic review and meta-analysis of randomised controlled trials and longitudinal observational studies. Risk of bias was assessed using ROB-2 and ROBINS I-V2.

**Data sources:** MEDLINE, Web of Science, EMBASE, and Cochrane Library to 27 March 2026.

**Eligibility criteria for selecting studies:** Studies which measured dementia or cognitive decline in women who initiated systemic MHT between ages 45-60 or within 5 years of menopause, compared with placebo or no MHT. Authors contacted for additional details if needed.

**Main outcome measures:** Dementia, Alzheimer’s disease (AD), cognitive decline.

**Results:** 10 studies totalling 213,678 participants (189,525 in studies with the primary population). There was no significant increased risk in women with a uterus for all cause dementia (pooled hazard ratio (HR): 1.12; 95% CI 0.91-1.31, N=78,613, I^2^ = 96.9%), but increased AD risk (HR: 1.14; 95% CI 1.02, 1.29, N=134,865, I^2^ = 35.6%). Results were similar in sensitivity analyses including women with or without a uterus. Results for cognitive decline were variable.

**Conclusions:** MHT initiated around the age of menopause should not be prescribed for cognition or dementia prevention. It is not protective against dementia and may increase risk slightly. The magnitude of risk was similar in AD and dementia, but the latter with larger confidence intervals. Studies which followed up individuals rather than on health records lost people to follow up. This may account for difference in cognitive decline outcomes between studies, as people with cognitive impairment and dementia are more likely not to attend.

MHT prescribing should balance benefits against risks, including evidence of a small increased dementia risk. There are few high-quality studies, so further research would inform recommendations.

**Systematic review registration** <u>Prospero CRD420251010663</u>

*What is already known on this topic?:*

1. Menopausal hormone therapy (MHT) is effective for alleviating vasomotor symptoms. Contemporary guidelines recommend treatment should be initiated for such symptoms under age 60 and or within 10 years of menopause onset.
2. A large randomised trial on the topic found increased risk of dementia in women initiating MHT after the age of 65.
3. It is unknown whether initiating MHT around the age of menopause impacts the risk of dementia or cognitive decline.

*What this study adds:*

1. There was no evidence that taking MHT around the time of menopause decreases the risk of dementia or cognitive impairment.
2. They should not be prescribed for these indications.
3. We were able to find more studies which examine this question by contacting authors for additional data.
4. Initiating MHT in women with a uterus around the age of menopause increased the risk of Alzheimer’s disease slightly, by over 10%, and there is a similar but not significant effect in the fewer studies of all cause dementia. Women with or without a uterus show similar results.
5. We found no significant difference shown in cognitive decline, possibly due to loss to follow up. This may be because most studies of cognitive decline follow up

## Introduction

Menopause occurs 12 months after the last menstrual period, and typically occurs naturally between 45 and 55 years old (median age 51).^1^ Around half of women experience vasomotor symptoms (hot flushes), as well as other symptoms like vulvovaginal atrophy (vaginal dryness or itching) and sleep disturbances, markedly impacting quality of life.^2,3^ Menopausal hormone therapy (MHT) alleviates vasomotor symptoms through oestrogen replacement.^4,5^ Additional progestogen (opposed MHT) is recommended for woman with a uterus to prevent oestrogen-induced endometrial malignant proliferation, whereas women without a uterus usually take unopposed (oestradiol-only) MHT.^6^ MHT was commonly prescribed from the 1990s until 2002, when the Women Health Initiative Study (WHIS) stopped their MHT randomised controlled trial (RCT) in women aged 50-79 due to physical health risk.^7^ However, MHT use increased again when it was found that it carried a better safety profile if started within ten years of menopause, as recommended by contemporary guidelines.^8^ The Women’s Health Initiative Memory Study (WHIMS), an adjunct RCT included women aged ≥ 65 years, found that those receiving MHT had twice the dementia risk over five years compared to those on placebo.^9^ Another study from the same trial found an increase in clinically important cognitive decline for those on MHT versus placebo.^10^

There is uncertainty about the cognitive consequences of taking MHT, with mixed findings on associated risks of cognitive deterioration and dementia. Two meta-analyses suggested a potential preventive effect of midlife MHT on dementia.^11,12^ Subsequently, a meta-analysis including three studies each of MHT initiated between 45-55 years old for opposed or unopposed MHT reported a non-significant increased risk of dementia and AD.^13^ Overall, meta-analyses have been contradictory, though there has been some suggestion that MHT could be prescribed in midlife to prevent dementia.

We aimed to clarify the impact of MHT use around the age at natural menopause (45-60, as recommended by contemporary guidelines) on future dementia and cognitive decline, initially in women with a uterus and then including those without to examine any difference.

## Methods

We registered the review on PROSPERO (Prospero ID: CRD420251010663) and followed the Preferred Reporting Items for Systematic reviews and Meta-Analyses (PRISMA) checklist.^14^

### Search strategy

We searched MEDLINE, Web of Science, EMBASE, and Cochrane CENTRAL from inception till 3 April 2025. We repeated the same search strategy on 27 March 2026 to screen any new published research since the initial screening. We used search terms related to MHT (or hormone replacement therapy), dementia, Alzheimer’s disease (AD), or cognitive impairment (Appendix A). We also searched the reference lists of included papers.

### Eligibility criteria

We included papers fulfilling all the following criteria:

- RCT or observational design.
- Peer-reviewed, published primary studies.
- Participants initiating MHT (oestrogen-only or combined oestrogen and progesterone) between ages 45-60 or within 5 years of menopause. The primary study population was women with a uterus only. We then included studies which had women with or without a uterus (that initiated MHT within 5 years of the removal of their uterus) for sensitivity analyses.
- Participants took MHT systemically (i.e. not local treatment with vaginal oestrogen).
- No stated contraindications to MHT (e.g. breast cancer).
- Participants (≥95%) took MHT for at least a year.
- Treatment groups were compared with placebo or no MHT.
- The outcome measure was dementia (all-cause dementia or AD, measured by odds ratio, risk ratio, or hazards ratio) or change in cognition (mean difference).

### Study selection

We uploaded search results into Covidence software, which automatically deduplicated the papers. Two reviewers, RL and AC, independently screened all the titles and abstracts. RL and AC then discussed and resolved any conflicts. RL and one of AC, AS, or UR independently reviewed the full text of the remaining papers. Reasons for exclusion were selected through a pre-coded list (Appendix B). RL and AC discussed and resolved all conflicts about exclusion. We emailed the authors of papers which lacked information required for a decision (e.g. age of MHT initiation). If authors replied with additional information, papers were further screened to see if they met inclusion criteria. We amended the protocol and registration inclusion criteria to include women without a uterus for sensitivity analyses, in which we re-screened the papers.

### Data extraction, risk of bias, and certainty assessment

We created a standard data extraction template (Appendix C). Two reviewers (RL and one of AC, AS, or UR) independently extracted data from the included studies and this was validated by a third reviewer (GL or NM). If studies stratified opposed and unopposed MHT, we assumed the opposed MHT would be women with a uterus. RL and either AC, AS, or UR independently used the Risk Of Bias In Non-randomized Studies-of Interventions, Version 2 (ROBINS I-V2)^15^ tool to assess all cohort studies and Risk of Bias 2 (RoB-2)^16^ tool for RCTs (Appendix D). We created a Microsoft form (Appendix E) to answer the questions from the risk of bias tools to categorise the answers into high, moderate, or low risk of bias. RL and the reviewer who conducted the assessment for the paper discussed discrepancies until they came to a consensus. RL and AC independently conducted a certainty assessment for dementia and AD separately using the GRADE approach^17,18^ (Appendix F) and discussed discrepancies until there was a consensus. The results were validated by a third reviewer (GL or NM).

### Analysis

We described included studies narratively (categorised by dementia or cognitive decline). The meta-analysis included results of a study that provided a re-analysis results excluding participants who initiated MHT after 60 years old.^19^ It also included two studies reporting odds ratios,^20,21^ which were interpreted as being equal to hazard ratios owing to their nested case-control designs.^22^

We conducted separate meta-analyses for dementia and AD using the random-effects DerSimonian and Laird model, which accounts for the variance within each study and between the studies.^23^ We performed logarithmic transformations on the ratios and confidence intervals before analysing through Metan, a statistical software on Stata v19.5 (Appendix G).^24^ We calculated heterogeneity using I^2^, representing the variation from clinical or methodological differences in the studies included.^25^ An I^2^ above 75 is interpreted as considerable heterogeneity, and results are less likely to be generalisable.^26^

We conducted post-hoc sensitivity analyses, one including studies of women without a uterus as well as those with a uterus, and another excluding studies scoring severe risk of bias on quality ratings. We created a funnel plot and conducted an Egger’s test to quantify for publication bias (p≤0.05 indicating significant publication bias), using Stata’s package for meta-analysis (Appendix H).^27^

### Patient and public involvement

This study was done because of paucity of evidence on MHT and dementia risk, clinical observations of women’s concerns regarding its potential preventive effects on dementia, and research recommendations identified by NICE. Patients and the public were not directly involved in the research question, design, or manuscript preparation; however, informal discussions with peri- and postmenopausal women informed contextual understanding.

## Results

The search identified 3,643 titles. After deduplication, we screened 2,364 papers using the title and abstract and then reviewed the full text of 184. We contacted 37 authors asking for further information on the age of MHT initiation or surgical menopause status. Ten replied^19,20,28–35^ (Appendix I), and the others were excluded. Five studies with this additional information fitted the criteria.^19,20,28–30^ An additional paper was found from the reference list of an included paper.^30^ Six papers including only women with a uterus^19–21,30,36,37^ and four papers with or without a uterus^28,29,38,39^ fulfilled inclusion criteria. Details on inclusion and exclusion of studies are displayed in figure 1.

### Study characteristics

Of the ten included papers, one was conducted in Denmark^19^, one in the UK^20^, one in Finland^21^, and seven in the United States.^28–30,36–39^ Their characteristics are shown in table 1 and their reported results in table 2.

**Table 1.**
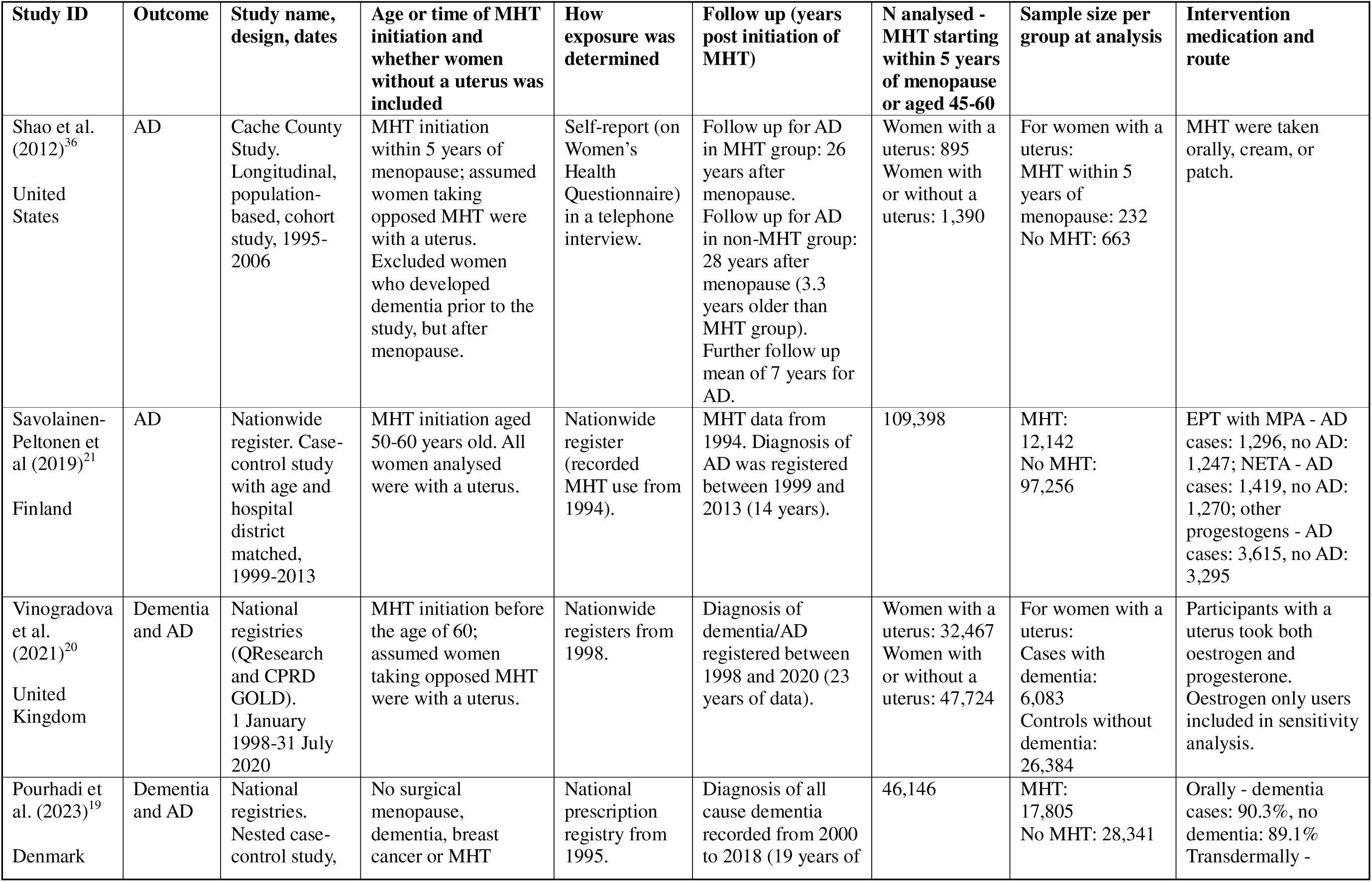
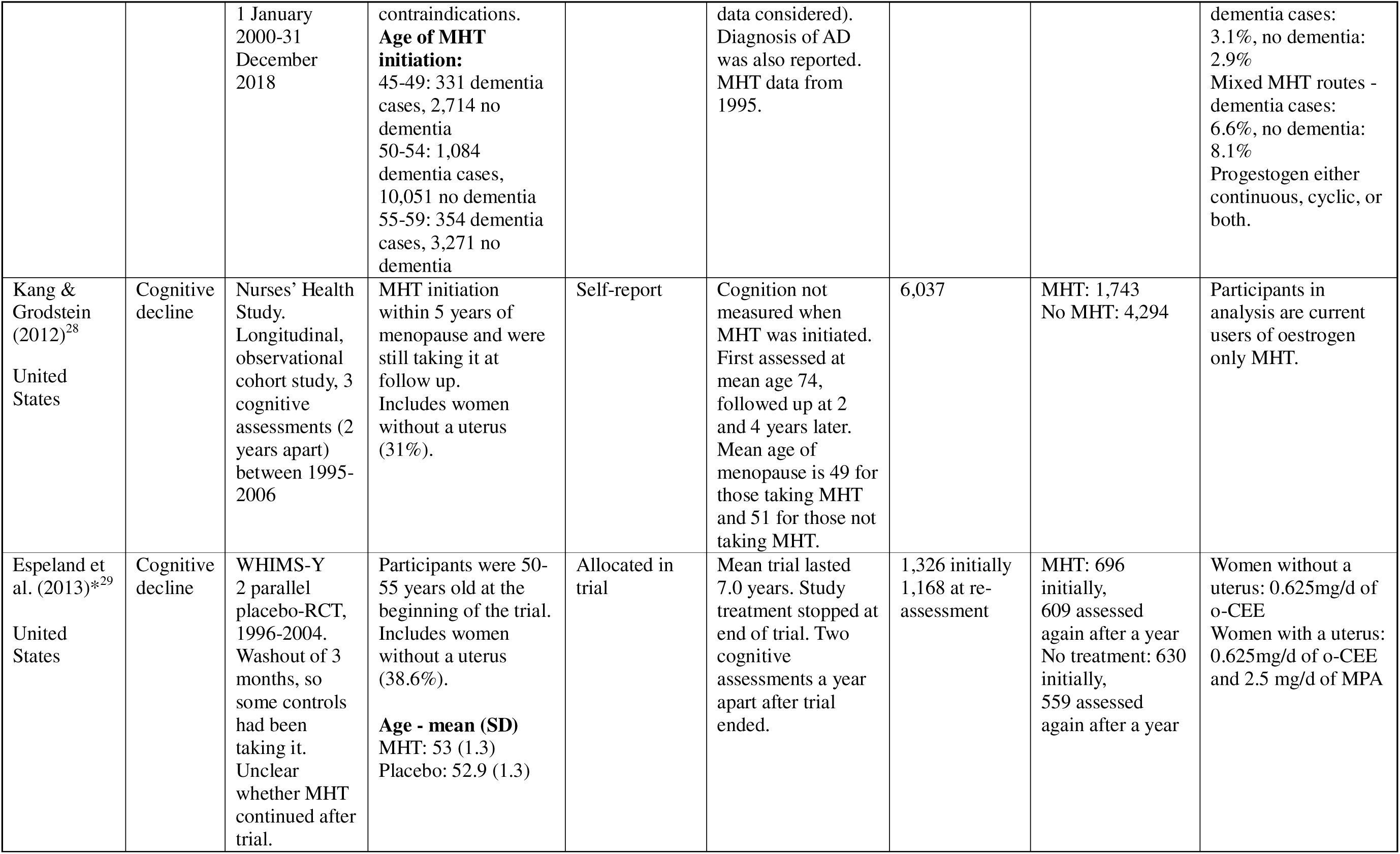
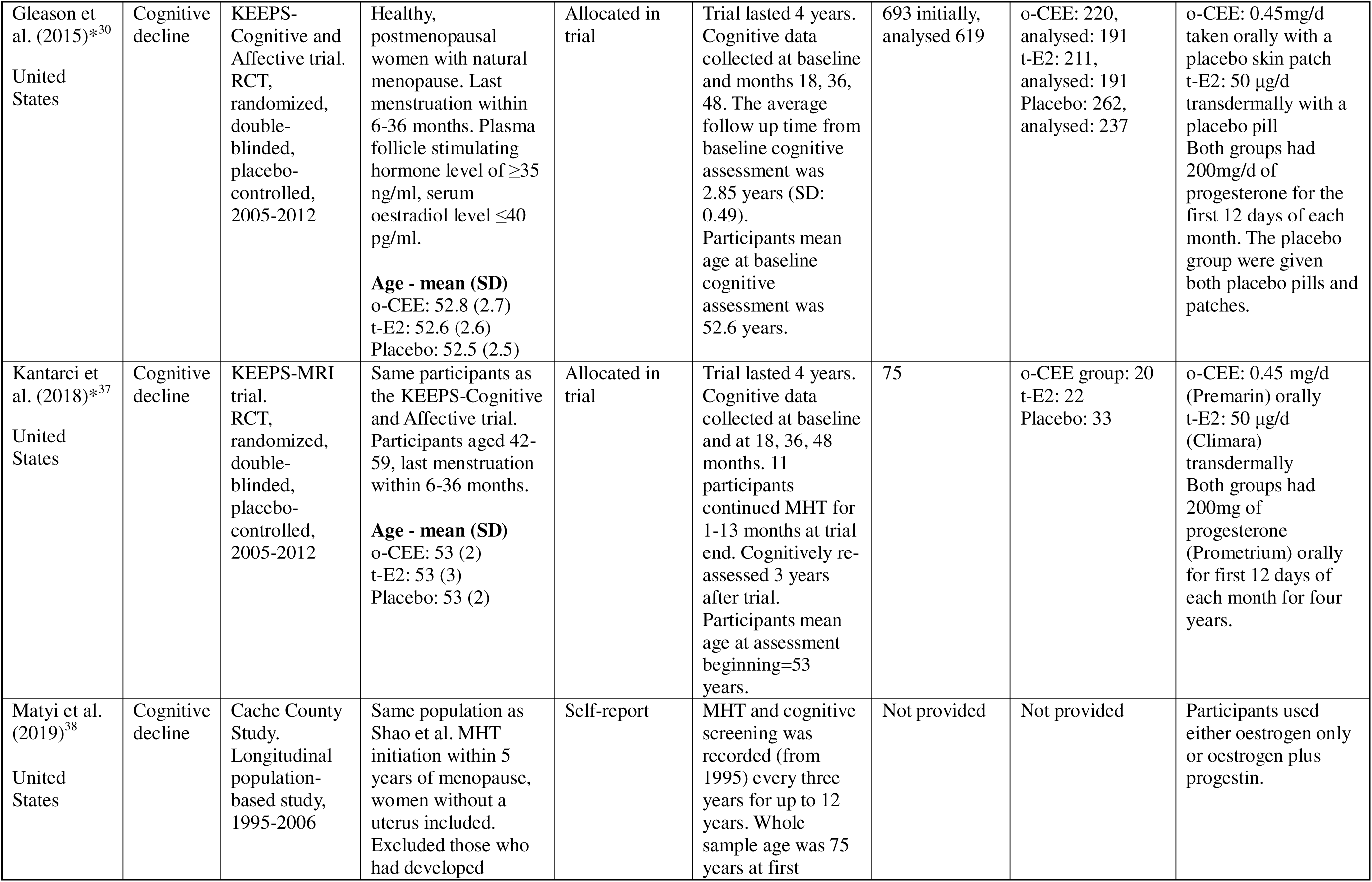
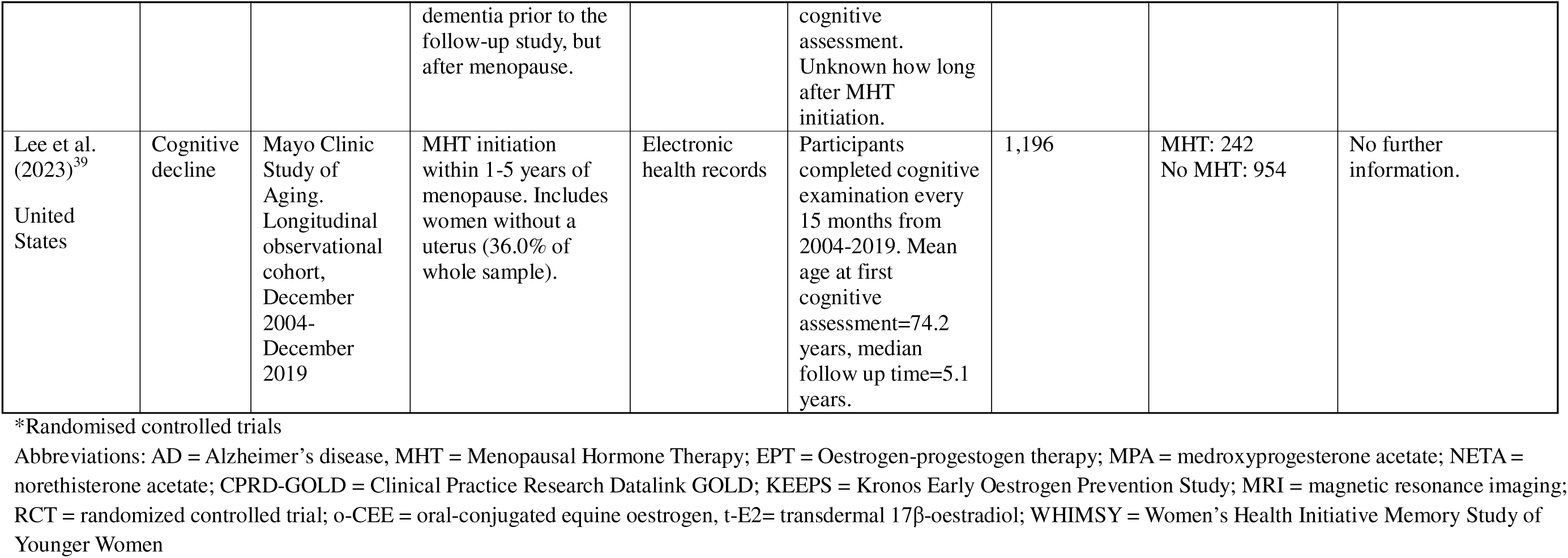
Characteristics of included papers.

| Study ID | Outcome | Study name, design, dates | Age or time of MHT initiation and whether women without a uterus was included | How exposure was determined | Follow up (years post initiation of MHT) | N analysed - MHT starting within 5 years of menopause or aged 45-60 | Sample size per group at analysis | Intervention medication and route |
| --- | --- | --- | --- | --- | --- | --- | --- | --- |
| Shao et al. (2012)<sup>36</sup><br>United States | AD | Cache County Study. Longitudinal, population-based, cohort study, 1995-2006 | MHT initiation within 5 years of menopause; assumed women taking opposed MHT were with a uterus. Excluded women who developed dementia prior to the study, but after menopause. | Self-report (on Women's Health Questionnaire) in a telephone interview. | Follow up for AD in MHT group: 26 years after menopause. Follow up for AD in non-MHT group: 28 years after menopause (3.3 years older than MHT group). Further follow up mean of 7 years for AD. | Women with a uterus: 895<br>Women with or without a uterus: 1,390 | For women with a uterus: MHT within 5 years of menopause: 232<br>No MHT: 663 | MHT were taken orally, cream, or patch. |
| Savolainen-Peltonen et al (2019)<sup>21</sup><br>Finland | AD | Nationwide register. Case-control study with age and hospital district matched, 1999-2013 | MHT initiation aged 50-60 years old. All women analysed were with a uterus. | Nationwide register (recorded MHT use from 1994). | MHT data from 1994. Diagnosis of AD was registered between 1999 and 2013 (14 years). | 109,398 | MHT: 12,142<br>No MHT: 97,256 | EPT with MPA - AD cases: 1,296, no AD: 1,247; NETA - AD cases: 1,419, no AD: 1,270; other progestogens - AD cases: 3,615, no AD: 3,295 |
| Vinogradova et al. (2021)<sup>20</sup><br>United Kingdom | Dementia and AD | National registries (QResearch and CPRD GOLD). 1 January 1998-31 July 2020 | MHT initiation before the age of 60; assumed women taking opposed MHT were with a uterus. | Nationwide registers from 1998. | Diagnosis of dementia/AD registered between 1998 and 2020 (23 years of data). | Women with a uterus: 32,467<br>Women with or without a uterus: 47,724 | For women with a uterus: Cases with dementia: 6,083<br>Controls without dementia: 26,384 | Participants with a uterus took both oestrogen and progesterone. Oestrogen only users included in sensitivity analysis. |
| Pourhadi et al. (2023)<sup>19</sup><br>Denmark | Dementia and AD | National registries. Nested case-control study, | No surgical menopause, dementia, breast cancer or MHT | National prescription registry from 1995. | Diagnosis of all cause dementia recorded from 2000 to 2018 (19 years of | 46,146 | MHT: 17,805<br>No MHT: 28,341 | Orally - dementia cases: 90.3%, no dementia: 89.1%<br>Transdermally - |
|  |  | 1 January 2000-31 December 2018 | contraindications.<br>**Age of MHT initiation:**<br>45-49: 331 dementia cases, 2,714 no dementia<br>50-54: 1,084 dementia cases, 10,051 no dementia<br>55-59: 354 dementia cases, 3,271 no dementia |  | data considered).<br>Diagnosis of AD was also reported.<br>MHT data from 1995. |  |  | dementia cases: 3.1%, no dementia: 2.9%<br>Mixed MHT routes - dementia cases: 6.6%, no dementia: 8.1%<br>Progestogen either continuous, cyclic, or both. |
| Kang & Grodstein (2012)<sup>28</sup><br>United States | Cognitive decline | Nurses' Health Study. Longitudinal, observational cohort study, 3 cognitive assessments (2 years apart) between 1995-2006 | MHT initiation within 5 years of menopause and were still taking it at follow up. Includes women without a uterus (31%). | Self-report | Cognition not measured when MHT was initiated. First assessed at mean age 74, followed up at 2 and 4 years later. Mean age of menopause is 49 for those taking MHT and 51 for those not taking MHT. | 6,037 | MHT: 1,743<br>No MHT: 4,294 | Participants in analysis are current users of oestrogen only MHT. |
| Espeland et al. (2013)<sup>29</sup><br>United States | Cognitive decline | WHIMS-Y 2 parallel placebo-RCT, 1996-2004. Washout of 3 months, so some controls had been taking it. Unclear whether MHT continued after trial. | Participants were 50-55 years old at the beginning of the trial. Includes women without a uterus (38.6%).<br>**Age - mean (SD)**<br>MHT: 53 (1.3)<br>Placebo: 52.9 (1.3) | Allocated in trial | Mean trial lasted 7.0 years. Study treatment stopped at end of trial. Two cognitive assessments a year apart after trial ended. | 1,326 initially<br>1,168 at re-assessment | MHT: 696 initially, 609 assessed again after a year<br>No treatment: 630 initially, 559 assessed again after a year | Women without a uterus: 0.625mg/d of o-CEE<br>Women with a uterus: 0.625mg/d of o-CEE and 2.5 mg/d of MPA |
| Gleason et al. (2015)*<sup>30</sup><br>United States | Cognitive decline | KEEPS-Cognitive and Affective trial. RCT, randomized, double-blinded, placebo-controlled, 2005-2012 | Healthy, postmenopausal women with natural menopause. Last menstruation within 6-36 months. Plasma follicle stimulating hormone level of $\geq 35$ ng/ml, serum oestradiol level $\leq 40$ pg/ml.<br>**Age - mean (SD)**<br>o-CEE: 52.8 (2.7)<br>t-E2: 52.6 (2.6)<br>Placebo: 52.5 (2.5) | Allocated in trial | Trial lasted 4 years. Cognitive data collected at baseline and months 18, 36, 48. The average follow up time from baseline cognitive assessment was 2.85 years (SD: 0.49). Participants mean age at baseline cognitive assessment was 52.6 years. | 693 initially, analysed 619 | o-CEE: 220, analysed: 191<br>t-E2: 211, analysed: 191<br>Placebo: 262, analysed: 237 | o-CEE: 0.45mg/d taken orally with a placebo skin patch<br>t-E2: 50 μg/d transdermally with a placebo pill<br>Both groups had 200mg/d of progesterone for the first 12 days of each month. The placebo group were given both placebo pills and patches. |
| Kantarci et al. (2018)*<sup>37</sup><br>United States | Cognitive decline | KEEPS-MRI trial. RCT, randomized, double-blinded, placebo-controlled, 2005-2012 | Same participants as the KEEPS-Cognitive and Affective trial. Participants aged 42-59, last menstruation within 6-36 months.<br>**Age - mean (SD)**<br>o-CEE: 53 (2)<br>t-E2: 53 (3)<br>Placebo: 53 (2) | Allocated in trial | Trial lasted 4 years. Cognitive data collected at baseline and at 18, 36, 48 months. 11 participants continued MHT for 1-13 months at trial end. Cognitively re-assessed 3 years after trial. Participants mean age at assessment beginning=53 years. | 75 | o-CEE group: 20<br>t-E2: 22<br>Placebo: 33 | o-CEE: 0.45 mg/d (Premarin) orally<br>t-E2: 50 μg/d (Climara) transdermally<br>Both groups had 200mg of progesterone (Prometrium) orally for first 12 days of each month for four years. |
| Matyi et al. (2019)<sup>38</sup><br>United States | Cognitive decline | Cache County Study. Longitudinal population-based study, 1995-2006 | Same population as Shao et al. MHT initiation within 5 years of menopause, women without a uterus included. Excluded those who had developed dementia prior to the follow-up study, but after menopause. | Self-report | MHT and cognitive screening was recorded (from 1995) every three years for up to 12 years. Whole sample age was 75 years at first cognitive assessment. Unknown how long after MHT initiation. | Not provided | Not provided | Participants used either oestrogen only or oestrogen plus progestin. |
| Lee et al. (2023)<sup>39</sup><br>United States | Cognitive decline | Mayo Clinic Study of Aging. Longitudinal observational cohort, December 2004-December 2019 | MHT initiation within 1-5 years of menopause. Includes women without a uterus (36.0% of whole sample). | Electronic health records | Participants completed cognitive examination every 15 months from 2004-2019. Mean age at first cognitive assessment=74.2 years, median follow up time=5.1 years. | 1,196 | MHT: 242<br>No MHT: 954 | No further information. |

\*Randomised controlled trials
Abbreviations: AD = Alzheimer's disease, MHT = Menopausal Hormone Therapy; EPT = Oestrogen-progestogen therapy; MPA = medroxyprogesterone acetate; NETA = norethisterone acetate; CPRD-GOLD = Clinical Practice Research Datalink GOLD; KEEPS = Kronos Early Oestrogen Prevention Study; MRI = magnetic resonance imaging; RCT = randomized controlled trial; o-CEE = oral-conjugated equine oestrogen, t-E2= transdermal 17β-oestradiol; WHIMSY = Women's Health Initiative Memory Study of Younger Women

**Table 2.** Results of included papers.

| Study ID | Outcome | Percentage followed up | Confounders | Primary outcomes | Secondary outcomes |
| --- | --- | --- | --- | --- | --- |
| Shao et al. (2012)<sup>36</sup> | AD (MMSE screen, then clinical assessment including informant interview). Only included attenders. | 1,769/2,090 (84.6%) FU. 321 had no final FU assessment, including 6 with known dementia at the first assessment they conducted. | Age, APOE status, years of education, predicted adherence to MHT use. | There was a non-significant reduction in AD risk for those initiating opposed MHT within 5 years of menopause (HR: 0.65, 95% CI 0.36, 1.18) compared to those who have not taken MHT. | Women who initiated unopposed MHT within 5 years of menopause showed a 35% significant decreased risk in AD compared to placebo. |
| Savolainen-Peltonen et al (2019)<sup>21</sup> | AD (from clinical records) | 100% as clinical databases. | Matched to controls by age and hospital district. | Initiating MHT before 60 years old was associated with an increased risk of AD (OR: 1.14, 95% CI 1.09, 1.19) compared to those who have not taken MHT. When looking at duration, increased risk of AD was not significant until after 5 years of taking MHT. | EPT combined with NETA and other progestogens had a significantly increased risk for dementia (OR: 1.17, 95% CI 1.08, 1.26; OR: 1.15, 95% CI 1.09, 1.21 respectively). EPT with MPA had a non-significant increased risk (OR: 1.08, 95% CI 1.00, 1.17).<br>Longer exposure to MHT was associated with a significant increased risk of AD (duration 5-10 years OR: 1.10, 95% CI 1.00, 1.20; 10 years or more OR: 1.20, 95% CI 1.13, 1.26). |
| Vinogradova et al. (2021)<sup>20</sup> | Dementia and AD (from clinical records) | 100% as clinical databases. | Smoking status, body mass index, family history of dementia, medical conditions and events, other medications, contraceptive drugs. | No significant difference in dementia risk in women initiating oestrogen and progesterone before 60 years old versus those who had not taken MHT (OR: 1.01, 95% CI 0.98, 1.04). Significant increased risk for AD in women initiating oestrogen and progesterone before 60 years old (OR: 1.13, 95% CI 1.08, 1.19). | When including women taking oestrogen only before the age of 60 into the sample, there was no difference in risk for dementia versus those who have not taken MHT (OR: 1.00, 95% CI 0.98, 1.03).<br>When including women taking oestrogen only before the age of 60 into the sample, there was a significant increased risk of AD versus those who have never taken MHT (OR: 1.10, 95% CI 1.05, 1.15). |
| Pourhadi et al. (2023)<sup>19</sup> | All cause dementia, late onset dementia, AD (from clinical records) | 100% as clinical databases. | Education, income, cohabitation, hypertension, diabetes, thyroid disease. | Initiating MHT from 45-60 led to an increased risk in developing all cause dementia (HR: 1.24, 95% CI 1.17, 1.33), late onset dementia (HR: 1.20, 95% CI 1.12, 1.30), and AD (HR: 1.22, 95% CI 1.07, 1.38). | **Treatment regimen**<br>Both continuous and cyclic treatment regimen increased the risk of dementia significantly.<br>**Duration (years)**<br>Longer exposure to MHT was associated with an increased risk for all cause dementia, late onset dementia, and Alzheimer's disease. |
| Kang & Grodstein (2012)<sup>28</sup> | TICS | 12,612/16,514 (76.4%) FU in overall study. | Age, education, diabetes, high blood pressure, vitamin E supplementation, body mass index, cigarette smoking, physical | Women who initiated oestrogen only MHT within five years of menopause and currently still taking them had a greater mean annual rate of cognitive decline than women who had never taken MHT ($\beta$: -0.05, 95% CI -0.10, -0.01). Those who had surgical menopause had | Surgical menopause group compared to MHT had similar results. No significant difference in specific cognitive domains. |
| | | | activity, socioeconomic status, antidepressant use, alcohol intake, aspirin use, nonsteroidal anti-inflammatory drug use, mental health index, energy-fatigue index. | annual decline $-0.12$ (95% CI $-0.23, -0.00$). | |
| Espeland et al. (2013)<sup>*29</sup> | TICS-m | 1,168/1,326 (88.1%) FU.<br>MHT: 609/696 (87.5%) FU.<br>Placebo: 559/630 (88.7%) FU. | Demographic, lifestyle, and clinical data related to risk of cognitive impairment, adherence. | No significant difference in TICS-m score in MHT group compared to placebo group ($\beta$: 0.02, 95% CI $-0.08, 0.12$). | No difference in specific cognitive domains measured for MHT group, aside from decrease in verbal fluency for the women without a uterus compared to placebo. |
| Gleason et al. (2015)<sup>*30</sup> | 3-MS | 518/693 (74.7%) FU.<br>MHT: 325/431 (75.4%) FU.<br>Placebo: 193/262 (73.7%) FU. | Baseline age, education. | No statistical difference in cognition for o-CEE ($\beta$: 0.01, 99% CI: $-0.00, 0.02$) or t-E2 ($\beta$: $-0.00, 99\%$ CI $-0.02, 0.01$) group versus placebo group. | No statistical difference in change for both treatment groups in specific cognitive domains measured. |
| Kantarci et al. (2018)<sup>*37</sup> | 3-MS | 75/101 (74.3%) had MRI and cognitive assessments at final follow up. | Groups balanced by randomisation. The mean age was same in each group. | No statistical difference in cognition for o-CEE ($\beta$: $-0.42, 95\%$ CI $-2.33, 1.48$) and t-E2 ($\beta$: $-0.08, 95\%$ CI $-1.91, 1.75$) group compared to placebo. | Significant increase in white matter hyperintensity and ventricular volumes in o-CEE group versus placebo at the end of the trial (48 months from baseline). No difference after last follow up assessment (3 years after trial ended). Dorso-prefrontal cortex volume was preserved for t-E2 group compared to placebo and associated with lower β-amyloid deposition. |
| Matyi et al. (2019)<sup>38</sup> | 3-MS | 2,114 at first wave.<br>No follow-up sample size provided. | Age, education, APOE genotype, body mass index, physical exercise, overall health, depression status. | Initiating MHT within 5 years of menopause had a significant positive association with cognitive score, scoring 1.23 points higher in women taking MHT compared to those not on MHT. Women with and without uterus were analysed together. | Initiating MHT within 1 year of menopause had a significant positive association with cognitive score ($\beta$: 1.02), while initiating MHT after 6 years or more were significantly negatively associated ($\beta$: 0.64). |
| Lee et al. | Global cognition | 2,193/2458 | Self-reported age, | No statistically significant difference in | Those initiating MHT within one year of |
| (2023)<sup>39</sup> | from memory, language, attention, visuospatial; domains MCI | (89.2%) FU. MHT within 1-5 years of menopause: 242/277 (87.4%) FU. Women not taking MHT: 954/1,056 (90.3%) FU. | education, hypertension, diabetes, dyslipidaemia, coronary heart disease, stroke, body mass index, smoking, APOE genotype. | cognitive decline in women initiating MHT within 1-5 years compared to those who had not ($\beta$: -0.01, 95% CI -0.03, 0.01). | menopause or earlier significantly improved in cognitive scores ($\beta$: 0.01, 95% 0.00, 0.03) compared to those who have not taken MHT, while those initiating more than five years after menopause did significantly worse ($\beta$: -0.04, 95% -0.05, -0.03). |

\* Randomised controlled trials
Abbreviations: MMSE = Mini Mental State Examination; FU = follow up; MHT = menopausal hormone therapy; TICS-m = Modified Telephone Interview for Cognitive Status; CI = confidence interval; AD = Alzheimer's Disease; HR = hazard ratio; CEE = conjugated equine oestrogen; 3-MS = Modified Mini Mental State; o-CEE = oral-conjugated equine oestrogen, t-E2= transdermal 17β-oestradiol; EPT = Oestrogen-progestogen therapy; MPA = medroxyprogesterone acetate; NETA = norethisterone acetate, MCI = mild cognitive impairment

Three papers analysed RCT data.^29,30,37^ Three studies were nationwide nested case-control studies,^19–21^ and four were prospectively gathered cohorts from population databases.^28,36,38,39^ Four studies’ outcome were dementia diagnoses (including AD),^19–21,36^ five measured cognitive decline,^28–30,37,38^ and one measured both mild cognitive impairment and cognitive decline.^39^

### Quality and certainty assessment

One study scored a low risk of bias,^30^ seven scored moderate risk,^19,20,28,36–39^ and two scored a high risk of bias^21,29^ (Appendix D). We found moderate certainty evidence that MHT does not protect from dementia and high certainty that MHT does not protect from AD (Appendix F).

### Main analysis

Four studies reported dementia or AD rates for women with a uterus initiating MHT within 5 years of menopause or age 45-60.^19–21,36^ Three of these used national registries with 19, 23 and 14 years of data considered (appendix J).^19–21^ The fourth was a longitudinal population-based cohort study.^36^ The study with 19 years of data included 46,146 women in total.^19^ Those initiating MHT from 45-60 years were more likely to develop all-cause dementia (HR: 1.24, 95% CI 1.17, 1.33) and AD (HR: 1.22, 95% CI 1.07, 1.38; N=11,812) than those not taking MHT. Longer MHT duration was associated with higher hazard ratios.^19^ The study with 23 years of data originally divided women into five groups according to treatment duration (Appendix K).^20^ We combined and meta-analysed among 32,467 women and found no significant increased risk of dementia (OR: 1.01, 95% CI 0.98, 1.04), but a significant increased risk for AD (OR: 1.13, 95% CI 1.08, 1.19; N=12,760).^20^ The third study with 14 years of data included 109,398 women and reported increased risk of AD among women taking MHT (OR: 1.14, 95% CI 1.09, 1.19).^21^ When stratified based on duration, this increased risk was significant after 5 years of taking MHT. The last included study looked at 895 women and found no significant risk of developing AD among women initiating MHT within 5 years of menopause compared to women who have not taken MHT (HR: 0.65, 95% CI 0.36, 1.18).^36^

In the meta-analysis, the overall pooled estimate from the two studies reporting dementia showed no significant risk in women with a uterus initiating MHT between 45-60 compared to those who have not taken MHT (HR: 1.12, 95% CI 0.91, 1.37, I^2^=96.9%; N=78,613, Figure 2).^19,20^ For AD, the pooled estimate of the four studies showed a significant increased risk in AD for women who initiated MHT around the age of natural menopause compared to women who have not taken MHT (HR: 1.14, 95% CI: 1.09, 1.20, I^2^=35.6%; N=134,865, Figure 3).^19–21,36^

### Including women without a uterus: AD

Two of the four papers also reported the associated AD risk when including women without a uterus. The study by Vinogradova et al reported no association with dementia in women initiating MHT before the age of 60 (OR: 1.00, 95% CI 0.98, 1.03; N=47,724) when they included women without a uterus (31.9% of sample).^20^ However, they found a significant increased risk of AD in women initiating MHT before the age of 60 (OR: 1.10, 95% CI 1.05, 1.15; N=18,527). Shao et al.’s paper showed a significant decreased risk of AD in women initiating MHT within 5 years of menopause (HR: 0.70, 95% CI 0.49, 0.99; N=1,390) when they included women without a uterus (23.7% of sample).^36^ When we add these figures into our meta-analysis, there was no statistically significant risk of dementia in women initiating MHT around the age of menopause compared to women who have not taken MHT (HR: 1.11, 95% CI 0.90, 1.37, I^2^=97.3%; N=93,870, Figure 4),^19,20^ but a significant increased risk of AD (HR: 1.12, 95% CI: 1.04, 1.20, I^2^=69.4%; N=142,075, Figure 5).^19–21,36^

We conducted a sensitivity analysis that excluded studies with serious risk of bias based on the ROBINS-I-V2 assessment. Three studies were included in this meta-analysis,^19,20,36^ and it continued to show a 14% increased risk of AD in women with a uterus initiating MHT around the age of menopause (HR: 1.14, 95% CI 1.01, 1.29, I^2^=57.0%; see fig 6). The funnel plot and Egger’s test indicated that there was no significant publication bias (p>0.05) for papers measuring risk of AD in women with a uterus initiating MHT around age of natural menopause (Appendix H).

### Cognitive decline

Six studies had an outcome of global cognitive decline, using a variety of cognitive tests. Three studies were RCTs,^29,30,37^ and three studies looked at longitudinal cohort studies.^28,38,39^ Three studies used the Modified Mini Mental State (3MS),^30,37,38^ two used the Telephone Interview for Cognitive Status or modified (TICS/TICS-m), ^28,29^ and one measured global cognition from average cognitive domains (memory, language, attention, visuospatial).^39^ Two studies had 7 years follow up^29,37^ and one with a mean of 2.8 years follow up.^30^ The remaining studies did not clearly state follow up time since initiation of MHT.^28,38,39^ *Cognitive decline: Women with a uterus*

One RCT divided women with a uterus aged 45-60 years into oral-conjugated equine oestrogen (o-CEE) and transdermal 17β-oestradiol (t-E2) groups.^30^ No significant difference was found in cognitive decline for both the o-CEE group (β: 0.01, 99% CI -0.00, 0.02, N=191 women analysed) and the t-E2 group (β: -0.00, 99% CI -0.02, 0.01, N=191 women analysed) compared to the control (N=237 women analysed) over a mean follow up of 2.85 years.

Kantarci and colleagues analysed the same trial, but with longer follow up and a smaller sample size.^37^ Both o-CEE (N=20) and t-E2 (N=22) showed no statistical difference compared to the placebo group (N=33) (β: -0.42, 95% CI -2.33, 1.48; β: -0.08, 95% CI -1.91, 1.75 respectively). However, a meta-analysis was not conducted because of insufficient information to allow for separating treatment from the placebo group.^30,37^

### Cognitive decline: Women with or without a uterus

There were an additional four studies which included participants with or without a uterus.^28,29,38,39^ We could not meta-analyse them due to different cognitive assessments, unknown confidence intervals,^38^ and outcome measuring standardised mean difference between placebo and treatment group.^29^ Two studies found no difference between MHT initiation around the age of menopause (or when surgical menopause was induced) compared to those who have not taken MHT.^29,39^ In one study, over half of the placebo group had previously used MHT and were randomised after a washout period (MHT N=609, placebo N=559 at follow up assessment).^29^ There was no significant difference observed (β: 0.02, 95% CI -0.08, 0.12). Another study found similar results when comparing 242 women initiating MHT within 1-5 years of menopause to 954 women who had never used MHT (β: -0.01, 95% CI -0.03, 0.01).^39^ A third non-randomised study found that women initiating MHT within five years of menopause improved in global cognition compared to those who had not taken MHT (β: 1.23, p=0.00).^38^ The last study found a significantly greater cognitive decline among 1,743 women who initiated oestrogen only MHT within five years of menopause, and were still using it at follow up assessment, compared with 4,294 women who has never taken MHT (β: -0.05, 95% CI -0.10, -0.01).^28^

## Discussion

We found MHT initiated near menopause onset is not protective against dementia, AD, or cognitive decline. It should not be prescribed to prevent them. There was a slightly increased risk for AD (14%) with a similar magnitude, but not statistically significant risk of all cause dementia in women. The AD analysis included more than twice as many women as all-cause dementia, suggesting a small, true effect with a lack of power. We found similar results including women with or without a uterus. The duration of MHT, if known, was associated with increasing risk.^19,21^ There was no increase in cognitive decline. In cohort studies, cognitively impaired individuals are more likely to be lost to follow-up, introducing potential attrition bias.^40^ Thus, the contradiction between results for cognitive decline and AD may be due to more cognitively impaired women being lost to follow-up,^28–30,36–39^ so it was less accurately estimated than dementia (which used routinely measured diagnosis).^19–21^

Our pre-registered study mainly included longitudinal, observational studies; only two RCTs met eligibility criteria. Where additional data were required, authors were contacted directly, enabling inclusion of a larger number of studies.^19,20,28–30^ Papers included used nationwide registers (N>32,000) or large-scale cohort studies (N=1,000-6,000), strengthening our analytic power.^19–21,28,36,38,39^ Our sensitivity analyses did not impact our results. Although natural menopause typically occurs in younger women, it can occur as late as age 60, so we used that as the upper age limit. A few, but not many participants, may therefore have initiated MHT up to 15 years after their final menstrual period, as most studies restricted MHT initiation to within five years of menstruation.^28,30,36–39^ As in correct clinical practice, we assumed oestrogen-only therapy was used by those who had a hysterectomy, but this may not always happen.^20,36^ Differences may have been attenuated in RCTs as some in the placebo group had taken MHT then had a washout period,^29,30,37^ potentially underestimating the true magnitude of effect. All included papers were from high income countries, with primarily white, healthy populations. In addition, hormone formulations have changed, so conjugated oestrogen is no longer used. Microionised progesterone and transdermal oestradiol are now standard, so present day prescriptions differ from older studies.^41^ This limits generalisability.^42^

Recent meta-analyses examining MHT initiation around menopause reported a non-significant increase in dementia risk for both oestradiol-only and combined MHT, consistent with our findings in magnitude and direction, but based on fewer studies.^13^ In contrast, a previous systematic review reported a 13.3% reduction in dementia risk associated with MHT use during midlife in observational studies, but did not define the age range used and selected oestrogen-only estimate in studies reporting both opposed and unopposed MHT, under-representing results showing increased dementia risk.^11^ A third systematic review found a 21.8% decreased risk of developing AD or dementia with any MHT use initiated at midlife (undefined) and included cross-sectional studies, which could not measure future risk.^12^ Our findings on cognitive decline align with those of an earlier systematic review of RCTs in which midlife initiation of MHT had no significant effect on global cognition scores relative to placebo.^43^

We do not know whether women with more severe menopausal symptoms, particularly brain fog, have an underlying predisposition to cognitive decline, though it does not present with memory impairment characteristic of AD and generally improved over time.^44^ The established association between MHT and vascular complications raises the possibility that this is a mechanism by which MHT could increase dementia risk, including AD.^7,45^ Further studies should examine outcomes by dementia subtype, including AD and vascular dementia separately, and should assess the effects of transdermal oestrogen and micronized progesterone in women initiating therapy close to the onset of natural menopause as is standard clinical practice.

## Data Availability

All data produced in the present study are contained in the manuscript.

## Acknowledgments

We would like to acknowledge the help of authors who gave us more information to enable us to use or exclude their papers.

## Transparency statement

The corresponding author affirms that the manuscript is an honest, accurate, and transparent account of the study being reported; that no important aspects of the study have been omitted; and that any discrepancies from the study as originally planned and registered have been explained.

## Contributors

GL and NP conceptualised the study. RL conducted the literature search and the initial calculations. RL, AC, AS, and UR screened the papers and data extraction. AC contributed to resolving the conflicts. RL wrote the first draft of the manuscript, and GL and NM checked the data extraction. All authors revised the paper for important intellectual content and agreed the final version. RL is the guarantor of the search. GL is the guarantor of the study and takes full responsibility for the content and conduct of the study and controlled the decision to publish.

The Corresponding Author has the right to grant on behalf of all authors and does grant on behalf of all authors, a worldwide licence to the Publishers and its licences in perpetuity, in all forms, formats and media (whether known now or created in the future), to i) publish, reproduce, distribute, display and store the Contribution, ii) translate the Contribution into other languages, create adaptations, reprints, include within collections and create summaries, extracts and/or, abstracts of the Contribution, iii) create any other derivative work(s) based on the Contribution, iv) to exploit all subsidiary rights in the Contribution, v) the inclusion of electronic links from the Contribution to third party material where—ever it may be located; and, vi) licence any third party to do any or all of the above.

## Declarations

There were no funding received and none of the authors had any competing interests. This study does not involve human participants, therefore no ethical approval was required.

---

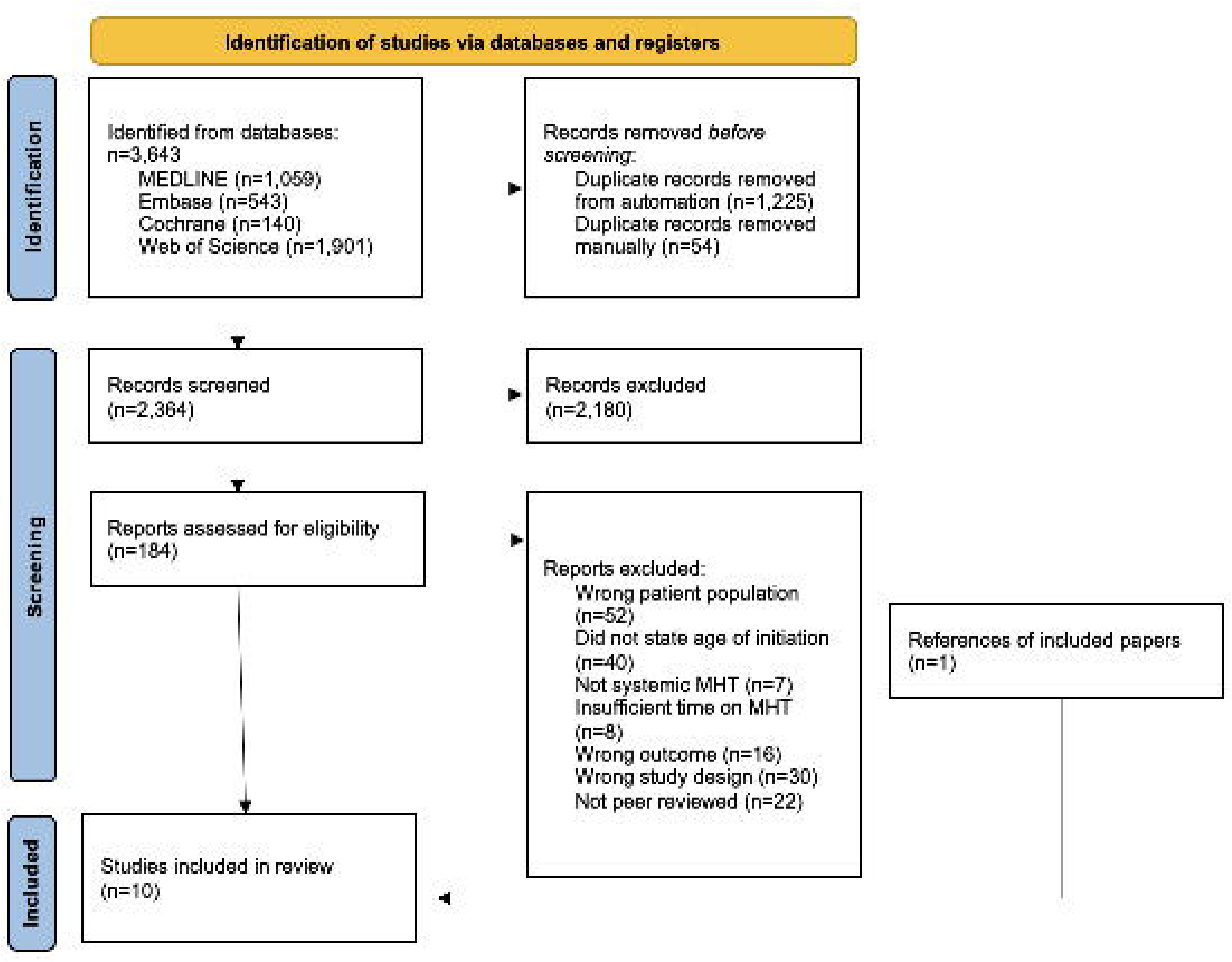

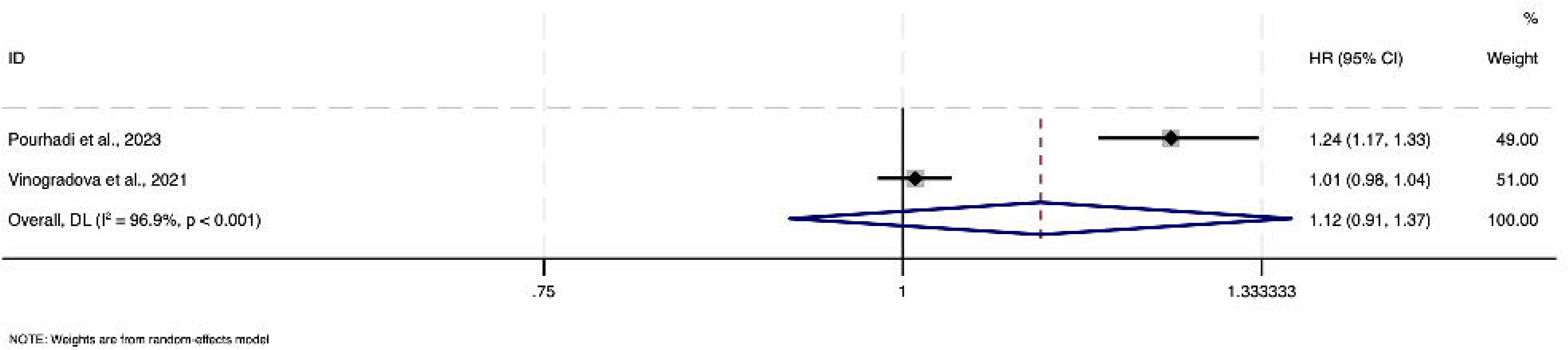

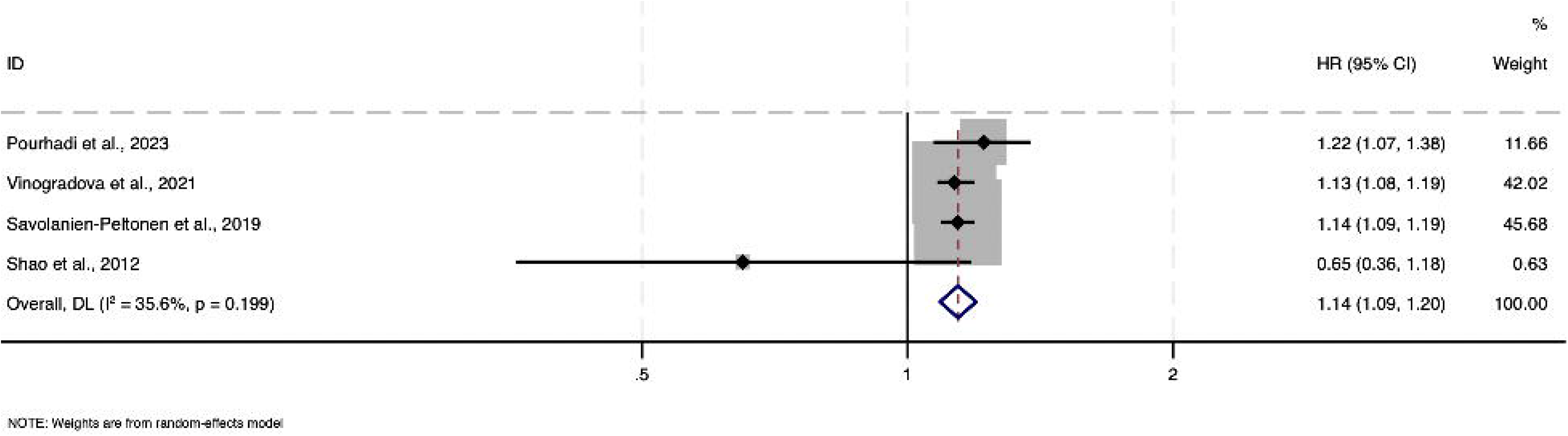

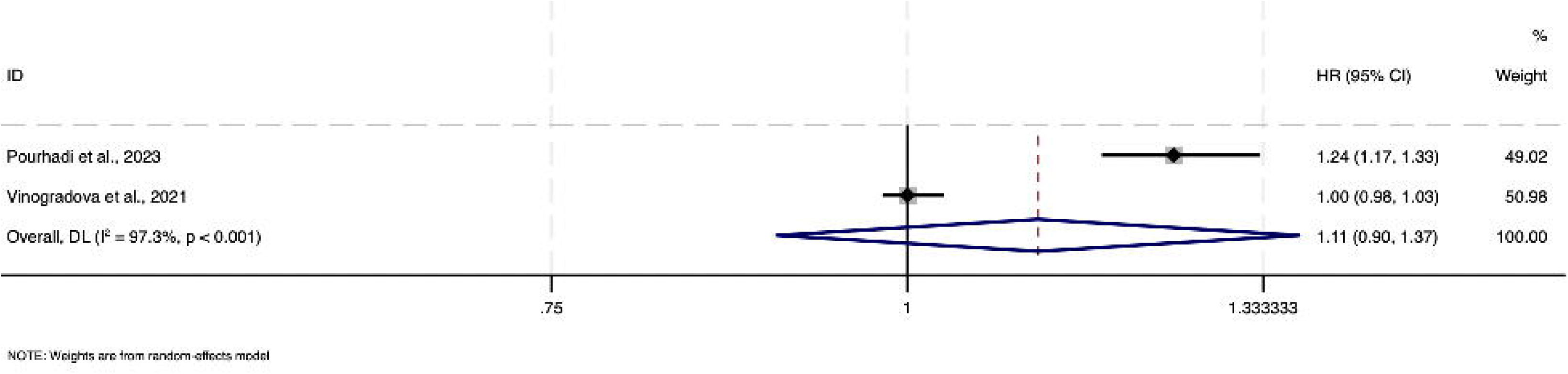

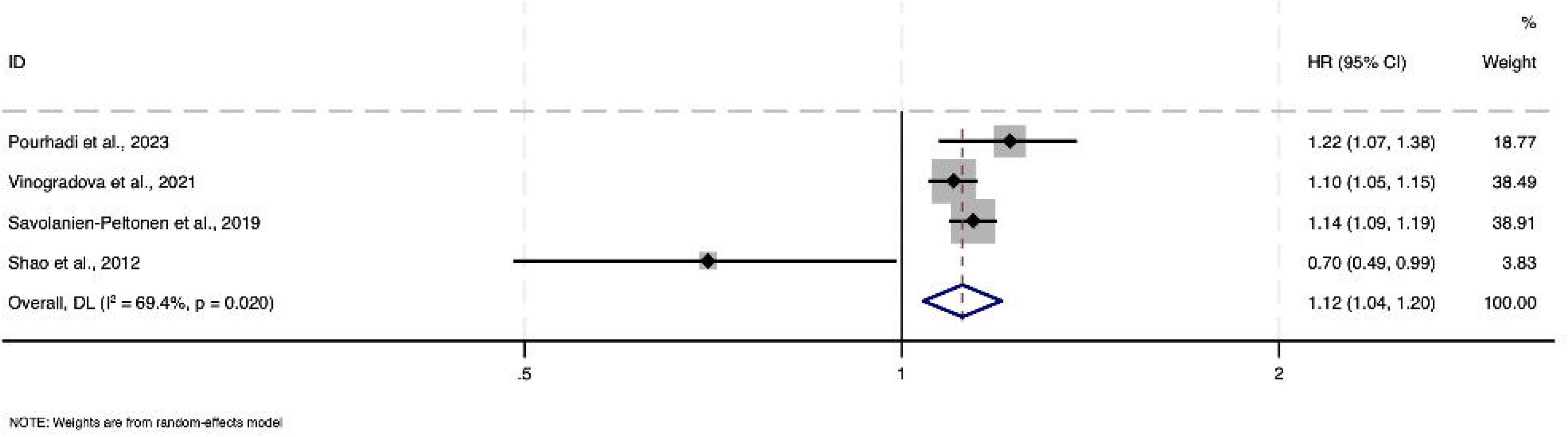

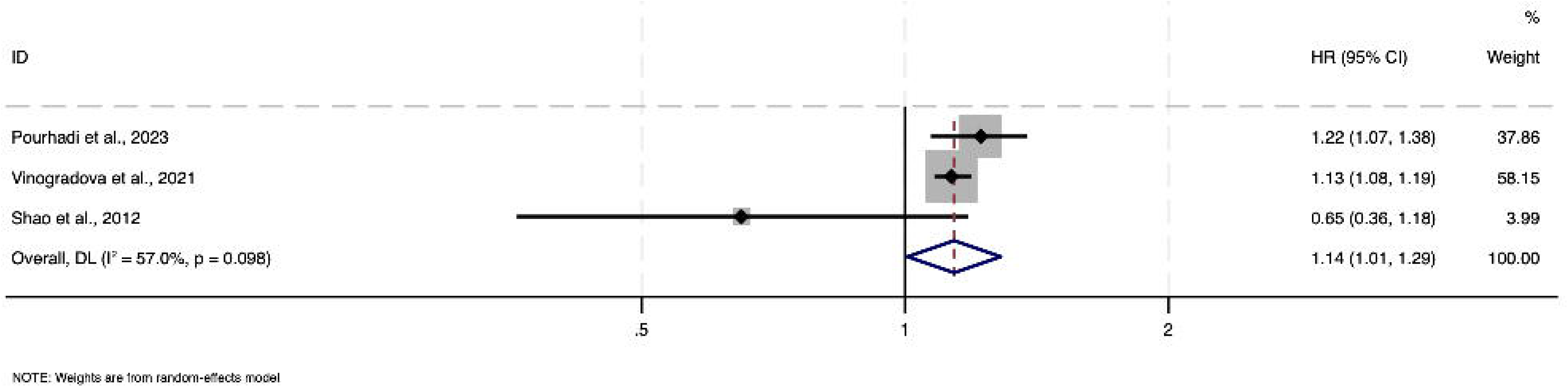

