## Appendix for "Menopausal hormone therapy at age 45 to 60 years old, future dementia or cognitive decline: Systematic review and meta-analysis"

**Appendix A**

‘Search strategy for each database’

| Database | MHT AND | Dementia |
| --- | --- | --- |
| MEDLINE | “Menopausal hormone therapy”* OR “hormone therapy” OR “oestrogen replacement therapy” OR “combined HRT” OR “oestrogen-only” OR “estrogen replacement therapy” OR “hormone replacement therapy” OR “menopausal hormone therapy” OR “hormone replacement therapy” | “dementia”* OR “dementia, multi-infarct” OR  “dementia” OR “frontotemporal dementia” OR “dementia, vascular” OR “Alzheimer's disease” OR “Lewy Body dementia” OR “frontotemporal dementia” OR “vascular dementia” OR “cognitive impair*” |
| EMBASE | “menopausal hormone therapy”* OR "hormone therapy" OR "oestrogen replacement therapy" OR "combined HRT" OR "oestrogen-only" OR “hormone substitution” OR “hormone therapy replacement” | “dementia”* OR “mixed dementia” OR “frontal variant frontotemporal dementia” OR “HIV associated dementia” OR “multiinfarct dementia” OR “presenile dementia” OR “semantic dementia” OR “frontotemporal dementia” OR “senile dementia” OR “Pick presenile dementia” OR “Alzheimer's disease” OR “Lewy Body dementia” OR “vascular dementia” OR “frontotemporal dementia” or “cognitive impair* |
| Cochrane | “Hormone replacement therapy”* OR "menopausal hormone therapy" OR “hormone therapy” OR "oestrogen replacement therapy" OR "combined HRT" OR "Oestrogen-only" | “Cognitive dysfunction” OR “dementia”* OR “Alzheimer’s disease” OR “Lewy Body dementia” OR “vascular dementia” OR “frontotemporal dementia” |
| Web of Science | “Menopausal hormone therapy”* OR “hormone replacement therapy” OR “hormone therapy” OR “oestrogen replacement therapy” OR “combined Hrt” OR “Hrt” OR “hormone replacement therapy Hrt” OR “estrogen replacement therapy” | “Dementia”* OR “vascular dementia” OR “frontotemporal dementia” OR “dementia with Lewy Bodies” OR “Alzheimer’s disease” OR “Lewy Body dementia” OR “Semantic dementia” or “people with dementia” or “cognitive impairment” OR “mild cognitive impairment” |

* MESH Term

We used the Boolean operator “OR” to separate each term and the * symbol as a Medical Subject Heading (MESH) term. We used the “AND” Boolean operators to combine MHT terms with dementia terms.

**Appendix B**

‘Pre-coded list of exclusion reasons’

- Wrong patient population e.g. MHT initiation not during 45-60 years
- Did not state age of initiation
- Not systemic MHT
- Insufficient time on MHT
- Wrong outcome
- Wrong study design
- Not peer reviewed

**Appendix C**

‘Data extraction table’

| **Study ID** | **Outcome** | **Study name, design, dates** | **Age or time of MHT initiation and whether surgical menopause included** | **How exposure was determined** | **Follow up (years post initiation of MHT)** | **N analysed– MHT starting within 5 years of menopause or aged 45-60** | **Sample size per group at analysis** | **Intervention medication and route** |
| --- | --- | --- | --- | --- | --- | --- | --- | --- |

| **Study ID** | **Outcome** | **Percentage followed up** | **Confounders** | **Primary outcomes** | **Secondary outcomes** |
| --- | --- | --- | --- | --- | --- |

**Appendix D**

‘Domains and results of quality assessment’

|  | 1 | 2 | 3 | 4 | 5 | 6 | 7 | Overall risk of bias |
| --- | --- | --- | --- | --- | --- | --- | --- | --- |
| Shao et al., 2012^36^~ |  |  |  |  |  |  |  | Moderate |
| Savolainen-Peltonen et al., 2019^21^~ |  |  |  |  |  |  |  | Serious |
| Vinogradova et al. 2021^20^~ |  |  |  |  |  |  |  | Moderate |
| Pourhadi et al., 2023^19^~ |  |  |  |  |  |  |  | Moderate |
| Kang & Grodstein, 2012^28^~ |  |  |  |  |  |  |  | Moderate |
| Espeland et al., 2013^29^* |  |  |  |  |  |  |  | Serious |
| Gleason et al., 2015^30^* |  |  |  |  |  |  |  | Low |
| Kantarci et al., 2018^37^* |  |  |  |  |  |  |  | Moderate |
| Matyi et al., 2019^38^~ |  |  |  |  |  |  |  | Moderate |
| Lee et al., 2023^39^~ |  |  |  |  |  |  |  | Moderate |

*RCTs assessed by RoB-2; ~Longitudinal studies assessed by ROBINS I-V2

Red = Serious, Yellow = Moderate, Green = Low

Savolainen-Peltonen et al. - serious risk of bias because they did not adjust for confounders, but mentioned that participant characteristics were most likely represented similarly in the case and control group.

Espeland et al. - serious risk of bias due to high percentage of missing data without specifying adjustments.

**ROBINS I-V2 domains:**

1) Bias from confounding, 2) bias in classification of interventions, 3) bias in selecting participants into the study (or analysis), 4) bias because of deviations from intended interventions, 5) bias from missing data, 6) bias from measurement of the outcome, 7) bias in selection of the reported results

**RoB-2 domains:**

1) Bias from randomization process, 2) bias because of deviations from intended interventions, 3) bias from missing outcome data, 4) bias in measurement of the outcome, 5) bias in selection of the reported results

**Appendix E**

‘Microsoft form for quality check assessments;

'Risk of Bias-2 Tool (ROB-2)’^16^

‘Risk Of Bias In Non-randomized Studies-of Interventions, Version 2’ (ROBINS-I-V2)^15^

**Risk of Bias-2 Tool (ROB-2)**: <https://forms.office.com/e/X2h92pBknz?origin=lprLink>

**‘Risk Of Bias In Non-randomized Studies-of Interventions, Version 2’ (ROBINS-I-V2)**:

<https://forms.office.com/e/nVu0WbWwMs?origin=lprLink>

**Appendix F**

‘GRADE approach to assess certainty of evidence’^17,18^

|  | **1** | **2** | **3** | **4** | **5** | **6** | **Total** |
| --- | --- | --- | --- | --- | --- | --- | --- |
| **Dementia** |  |  |  |  |  |  |  |
| **AD** |  |  |  |  |  |  |  |

Red = Serious, Yellow = Moderate, Green = Low

1) Risk of bias, 2) Imprecision, 3) Indirectness, 4) Inconsistency, 5) Publication bias, 6) Dose response effect

**Appendix G**

‘METAN code for dementia meta-analyses’^24^

 ssc install metan

gen lnHR = ln(HR)

gen lnuci = ln(upperCI)

gen lnlci = ln(bottomCI)

metan lnHR lnlci lnuci, eform effect(HR) random lcols(author)

meta set lnHR lnlci lnuci, random(dlaird) studylabel(author) civartolerance(.1)

meta summarize, eform random(dlaird)

metafunnel lnHR lnlci lnuci, eform

meta bias, egger

**Appendix H**

‘Funnel plot and Egger’s test for meta-analysis on women with a uterus and AD’^19–21,36^

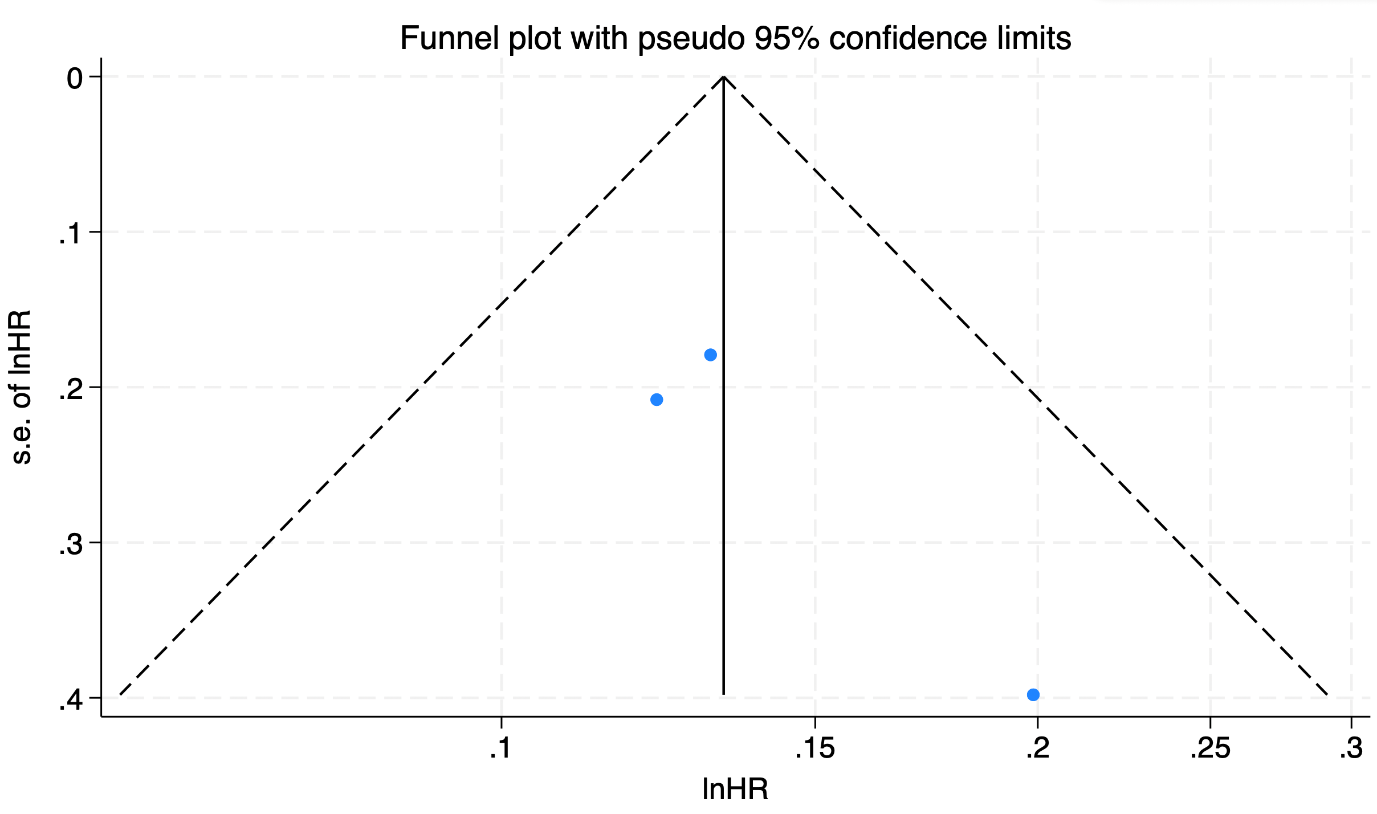

Egger’s test

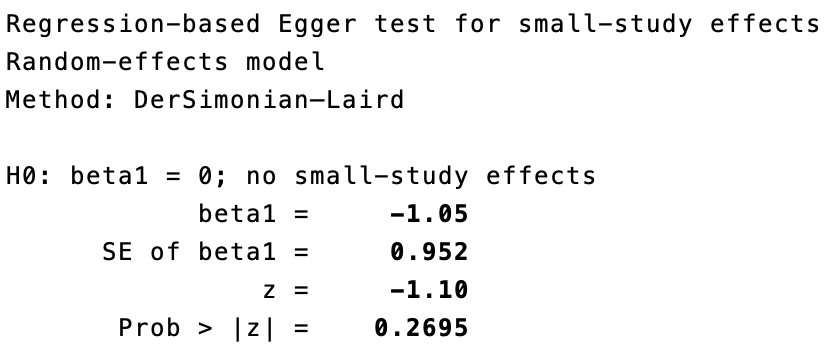

**Appendix I**

‘Papers and extra information from authors’

- Pourhadi N, Mørch LS, Holm EA, Torp-Pedersen C, Meaidi A. (2023) Menopausal hormone therapy and dementia: nationwide, nested case-control study.^19^
  - Provided reanalysed figures to exclude people initiating MHT >60 years as follows.
    - **Dementia:** Oestrogen-progestin, <60 at initiation (1769 cases): HR 1,24 95% CI (1.17-1.33) p<0.001; Oestrogen-progestin, ≥60 at initiation (13 cases): HR 1,24 95% CI (0.69-2.21) p<0.477
    - **Alzheimer's disease:** Oestrogen-progestin, <60 at initiation (471 cases, 4220 controls): HR 1,22 95% CI (1.07-1.38) p=0.003; Oestrogen-progestin, ≥60 at initiation (5 cases, 34 controls): HR 1,49 95% CI (0.57-3.90) p=0.417
    - **Late onset dementia:** Oestrogen-progestin, <60 at initiation (1390 cases, 12,677 controls): HR 1,20 95% CI (1.12-1.30) p<0.001; Oestrogen-progestin, ≥60 at initiation (13 cases, 98 controls): HR 1,40 95% CI (0.78-2.53) p=0.256
- Vinogradova Y, Dening T, Hippisley-Cox J, Taylor L, Moore M, Coupland C. (2021) Use of menopausal hormone therapy and risk of dementia: nested case-control studies using QResearch and CPRD databases.^20^
  - Included as provided information on proportion of women initiating MHT <45 years (2% for CPRD Gold and unsure for QResearch).
- Kang JH, Grodstein F. (2012) Postmenopausal hormone therapy, timing of initiation, APOE and cognitive decline.^28^
  - Included in the sensitivity analysis for cognitive decline since they included a portion of women without a uterus.
- Espeland MA. (2013) Long-term effects on cognitive function of postmenopausal hormone therapy prescribed to women aged 50 to 55 years.^29^
  - Included in the sensitivity analysis for cognitive decline since they included a portion of women without a uterus.
- Gleason CE, Dowling NM, Wharton W, Manson JE, Miller VM, Atwood CS, et al. (2015) Effects of hormone therapy on cognition and mood in recently postmenopausal women: findings from the randomized, controlled KEEPS–Cognitive and Affective study.^30^
  - Although paper included women with last menstrual period within 6 months, in answer to our queries they confirmed that they had checked that the oestradiol and FSH levels had menopausal status.
- Lokken KL, Ferraro FR. (2006) The relationship between menopausal status, phase of menstrual cycle, and replacement estrogen on cognition in healthy women without dementia.^31^
  - Excluded because they did not collect age of MHT initiation.
- Seshadri S, Zornberg GL, Derby LE, Myers MW, Jick H, Drachman DA. (2001) Postmenopausal estrogen replacement therapy and the risk of Alzheimer Disease.^32^
  - Excluded because they did not collect age of MHT initiation.
- Levine AJ, Hewett L. (2003) Estrogen replacement therapy and frontotemporal dementia.^33^
  - Excluded as they did not collect age of MHT initiation.
- Fillenbaum GG, Hanlon JT, Landerman LR, Schmader KE. (2001) Impact of estrogen use on decline in cognitive function in a representative sample of older community-resident Women.^34^
  - Excluded because they did not state the age of initiation for MHT.
- Barrett-Connor E, Kritz-Silverstein D. (1993) Estrogen replacement therapy and cognitive function in older women.^35^
  - Provided the Rancho Bernado Study open access database to individually analyse. After attempting to analyse, we found it was not possible due to repeated data of the same participant with different results and missing data.

**Appendix J**

‘Hospital registers used’

- Pourhadi N, Mørch LS, Holm EA, Torp-Pedersen C, Meaidi A. (2023) Menopausal hormone therapy and dementia: nationwide, nested case-control study.^19^
  - Danish National Prescription Registry for MHT use
  - Danish National Registry of Patients for dementia
- Vinogradova Y, Dening T, Hippisley-Cox J, Taylor L, Moore M, Coupland C. (2021) Use of menopausal hormone therapy and risk of dementia: nested case-control studies using QResearch and CPRD databases.^20^
  - QResearch - general practice records, hospital episode statistics, mortality data to identify cases and controls
  - CPRD GOLD - cases from general practice records
- Savolainen-Peltonen H, Rahkola-Soisalo P, Hoti F, Vattulainen P, Gissler M, Ylikorkala O, et al. (2019) Use of postmenopausal hormone therapy and risk of Alzheimer’s disease in Finland: nationwide case-control study^21^
  - Finnish drug reimbursement register for cases.
  - Finnish National Population Register for age match controls.

**Appendix K**

‘Meta-analyses results from Vinogradova’s study’^20^

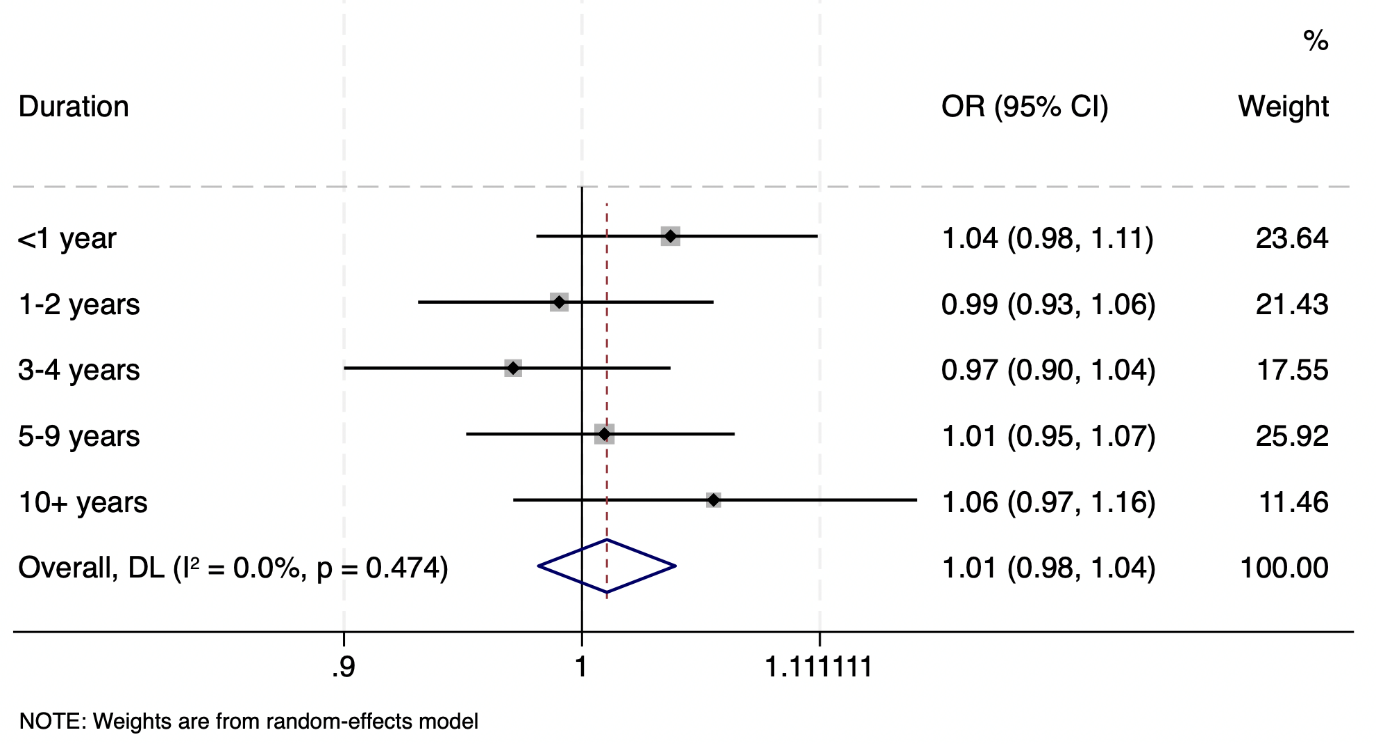
Risk for dementia in women initiating oestrogen and progesterone before 60 years old.

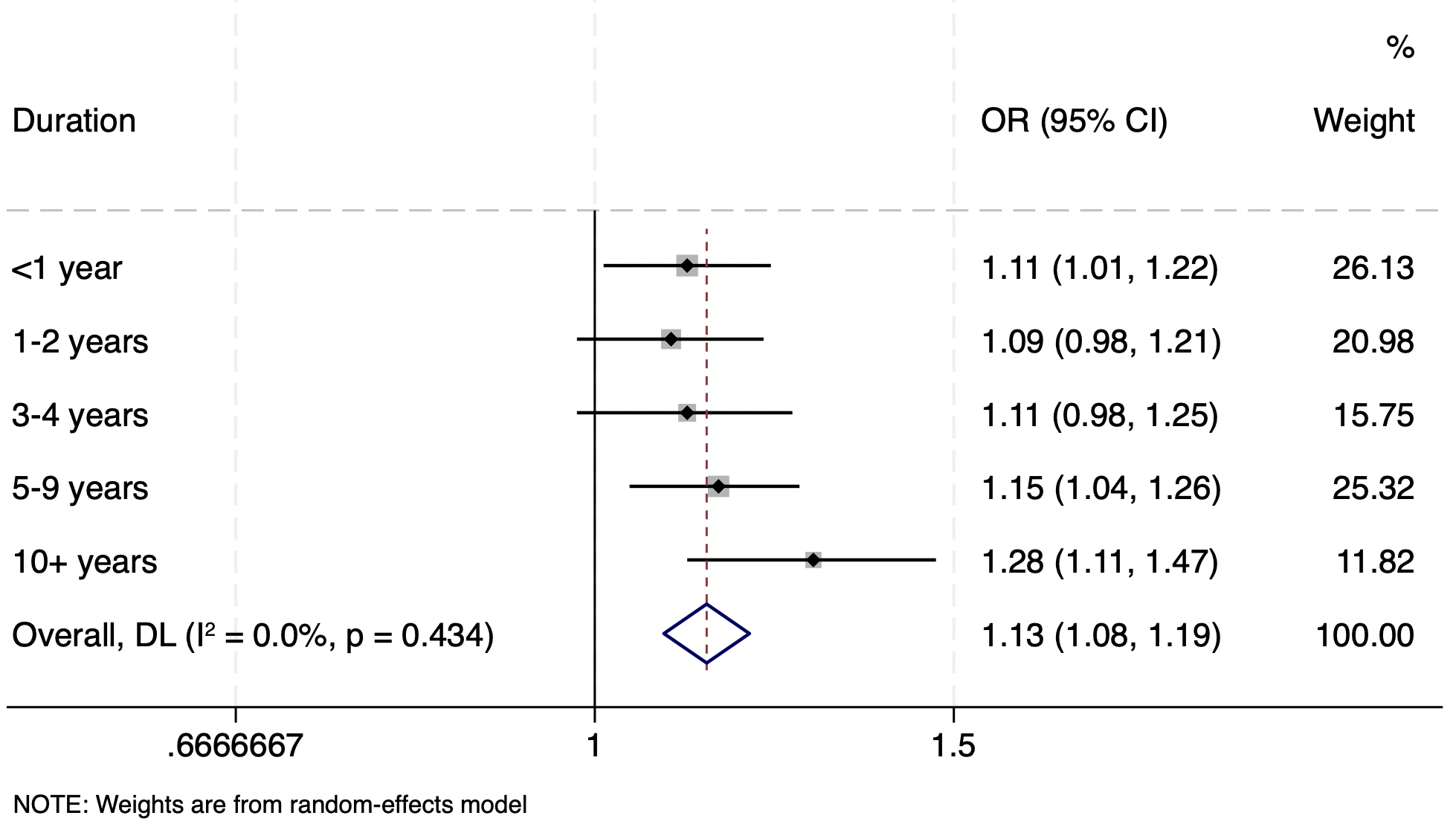

Risk for AD in women initiating oestrogen and progesterone before 60 years old

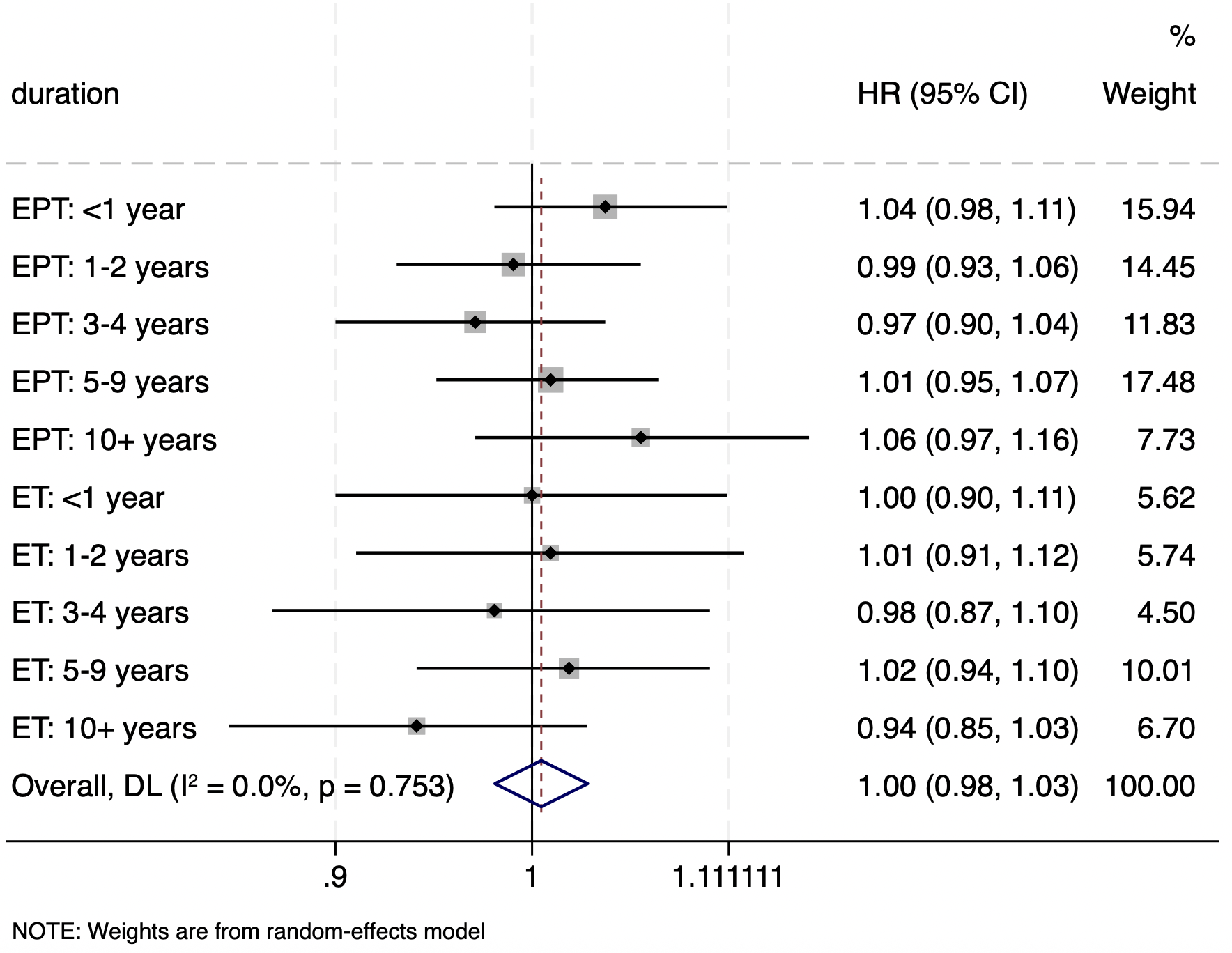

Risk for dementia in women initiating either oestrogen-only or oestrogen and progesterone before the age of 60
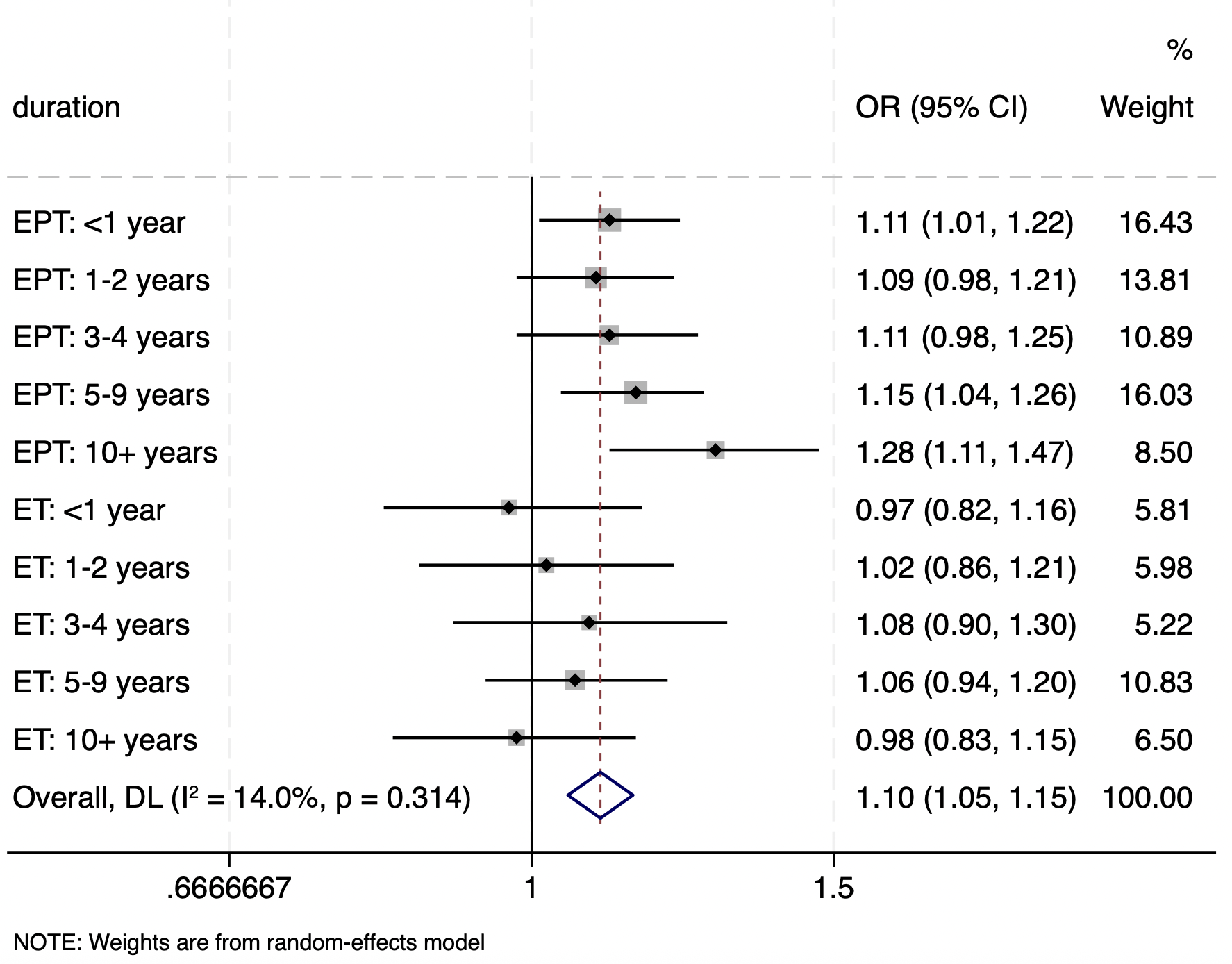

Risk for AD in women initiating either oestrogen-only or oestrogen and progesterone before the age of 60
